# Evaluating the Use of GLP-1 Receptor Agonists in Wolfram syndrome Patients

**DOI:** 10.64898/2026.03.31.26349885

**Authors:** Laura Lee, Abby F. Tang, Anna Asako, Sarah F. Ning, Hayden A. Reed, Eleanor Duncan, Heather M. Lugar, James Hoekel, Bess A. Marshall, Tamara Hershey, Fumihiko Urano

**Affiliations:** Department of Medicine, Division of Endocrinology, Metabolism, and Lipid Research, Washington University School of Medicine, 660 South Euclid Avenue, St. Louis, MO 63110, USA; Department of Psychiatry, Washington University School of Medicine, 660 South Euclid Avenue, St. Louis, MO 63110, USA; Department of Ophthalmology & Visual Sciences, Washington University School of Medicine, 660 South Euclid Avenue, St. Louis, MO 63110, USA; Departments of Pediatrics and Cell Biology, Division of Endocrinology and Diabetes, Washington University School of Medicine, 660 South Euclid Avenue, St. Louis, MO 63110, USA; Department of Neurology, Washington University School of Medicine, 660 South Euclid Avenue, St. Louis, MO 63110, USA; Department of Radiology, Washington University School of Medicine, 660 South Euclid Avenue, St. Louis, MO 63110, USA; Department of Pathology and Immunology, Washington University School of Medicine, 660 South Euclid Avenue, St. Louis, MO 63110, USA

**Keywords:** Wolfram syndrome, WFS1, GLP-1 receptor agonists, diabetes mellitus, optic atrophy, endoplasmic reticulum stress, incretin therapy, rare disease

## Abstract

Wolfram syndrome is a rare autosomal recessive disorder caused by pathogenic variants in the *WFS1* gene, characterized by early-onset diabetes mellitus, optic atrophy, sensorineural hearing loss, arginine vasopressin deficiency, and progressive neurodegeneration. The condition selectively affects pancreatic β cells and neurons via chronic endoplasmic reticulum (ER) stress, and no proven disease-modifying therapy currently exists. Diabetes mellitus is typically the first manifestation, presenting at a mean age of 6 years as an insulin-dependent phenotype with preserved C-peptide and negative diabetes-related autoantibodies.

Glucagon-like peptide-1 receptor agonists (GLP-1 RAs) are well-established agents in the management of type 2 diabetes, augmenting glucose-dependent insulin secretion, suppressing glucagon, slowing gastric emptying, and promoting satiety. Preclinical evidence further suggests that GLP-1 RAs preserve β-cell mass, attenuate ER stress, and confer neuroprotective effects, properties of particular therapeutic relevance to Wolfram syndrome.

We conducted a retrospective cohort study of 84 participants with genetically confirmed Wolfram syndrome and insulin-dependent diabetes mellitus enrolled in the Washington University Wolfram Syndrome International Registry and Clinical Study. Clinical data were extracted from medical records; for participants concurrently enrolled in the Tracking Neurodegeneration in Early Wolfram Syndrome study, longitudinal data were obtained from that source as well. Thirty-five percent of eligible participants had received a GLP-1 RA at some point during follow-up. We characterize the prevalence of GLP-1 RA use, documented rationale for initiation, observed effects on glycemic control and visual outcomes, adverse effects, and reasons for discontinuation. No statistically significant changes in hemoglobin A1c (HbA1c) or body mass index (BMI) were observed. Visual acuity declined significantly at two years, consistent with expected disease progression. Gastrointestinal adverse effects were common and contributed to frequent discontinuation.

These observational data provide important clinical context and a foundation for future prospective trials evaluating GLP-1 RAs as a potential disease-modifying strategy in Wolfram syndrome.

## Introduction

Wolfram syndrome is a rare autosomal recessive disorder with a prevalence of approximately 1 in 250,000 to 700,000 individuals (Urano, 2016;Mishra et al., 2021;Lee et al., 2023). It is caused by biallelic pathogenic variants in the *WFS1* gene, which encodes wolframin, an 890-amino acid transmembrane protein localized to the endoplasmic reticulum (ER) and implicated in calcium homeostasis and protein quality control (Inoue et al., 1998;Fonseca et al., 2005;Fonseca et al., 2010;Lu et al., 2014). Loss of function of *WFS1* leads to dysregulated ER stress signaling, particularly in pancreatic β cells and neurons, which are especially vulnerable to chronic ER stress-mediated apoptosis (Fonseca et al., 2010).

The classic clinical symptoms of Wolfram syndrome encompasses diabetes mellitus, optic nerve atrophy, sensorineural hearing loss, arginine vasopressin deficiency (previously termed diabetes insipidus), and progressive neurodegeneration (Barrett et al., 1995;Hershey et al., 2012). Diabetes mellitus is typically the earliest manifestation, diagnosed at a mean age of approximately 6 years, and is distinguished by positive C-peptide levels and absence of autoantibodies which are markers of type 1 diabetes, reflecting progressive ER stress-driven β cell dysfunction rather than autoimmune destruction (Ray et al., 2022;Lee et al., 2023). Optic nerve atrophy typically follows in the second decade, with a mean onset around 11 years (O’Bryhim et al., 2022;Lee et al., 2023;McNeely et al., 2026). The mortality rate exceeds 50% by age 40 years, with respiratory failure due to complications of brainstem atrophy (Kinsley et al., 1995). No proven disease-modifying therapy currently exists for Wolfram syndrome (Abreu and Urano, 2019).

GLP-1 receptor agonists (GLP-1 RAs) are a class of medications that mimic endogenous glucagon-like peptide-1, an incretin hormone expressed widely in the gut and central nervous system (Drucker, 2025b;a;Moiz et al., 2025;Drucker, 2026;Nauck et al., 2026). In the pancreas, GLP-1 RAs enhance glucose-dependent insulin secretion and suppress glucagon release; peripherally, they slow gastric emptying and increase satiety. Beyond these metabolic effects, GLP-1 RAs promote neuronal differentiation, inhibit neuroinflammation, reduce ER stress, and exert beneficial effects on vascular endothelium (Cheng et al., 2022;Kopp et al., 2022;Chen et al., 2024;Gong et al., 2025). Those are properties of particular mechanistic relevance to Wolfram syndrome.

Preclinical evidence provides a compelling rationale for GLP-1 RA use in Wolfram syndrome. It has been demonstrated that 4 weeks of exendin-4 treatment in *Wfs1* knockout mice restored glucose-stimulated insulin secretion and alleviated β cell ER stress (Kondo et al., 2018). It has been also shown that early and sustained GLP-1 RA therapy delayed the onset of diabetes and visual neurodegeneration in Wolfram syndrome rodent models (Plaas et al., 2017;Toots et al., 2018;Seppa et al., 2019;Jagomae et al., 2021). In human models, GLP-1 RAs improved β cell function and prevented apoptosis in *WFS1*-deficient induced pluripotent stem cell-derived β cells and neurons (Gorgogietas et al., 2023). A few case reports have described off-label GLP-1 RA use in Wolfram syndrome patients over observation periods of 8 to 27 months, with authors noting an absence of measurable neuro-ophthalmological progression during the treatment period and reporting generally good tolerability with transient mild gastrointestinal side effects. As with any uncontrolled case series, the absence of placebo controls and the relatively limited follow-up periods make it difficult to determine whether the observed stability reflects a true treatment effect or the natural variability in disease progression over short observation windows (Frontino et al., 2021;Panfili et al., 2023).

Based on this preclinical and limited clinical evidence, GLP-1 RAs have been increasingly adopted in clinical practice for Wolfram syndrome patients. Here, we report a retrospective cohort analysis of GLP-1 RA use in 84 participants with genetically confirmed Wolfram syndrome. We describe the prevalence of use, rationale for initiation, effects on glycemic control, body mass index, and visual parameters, adverse effects, and reasons for discontinuation.

## Materials and Methods

### Patients

Participants were selected from the Wolfram syndrome International Registry and Clinical Study. Clinical data were obtained from medical records and phone interviews; for participants who had also enrolled in the Tracking Neurodegeneration in Early Wolfram syndrome study, longitudinal data were additionally extracted from that source. Participants and their parents or legal guardians as appropriate, provided written informed consent before participating in this study, which was approved by the Human Research Protection Office at Washington University School of Medicine in St. Louis, MO. Participants were selected for inclusion based on: (1) two confirmed biallelic disease-causing *WFS1* variants on genetic testing; (2) insulin-dependent diabetes mellitus and (3) complete outcome measures data (see Clinical Variables section) available through electronic or paper medical records. 84 participants were included in the final analytic cohort (Figure 1).

**Figure 1.**
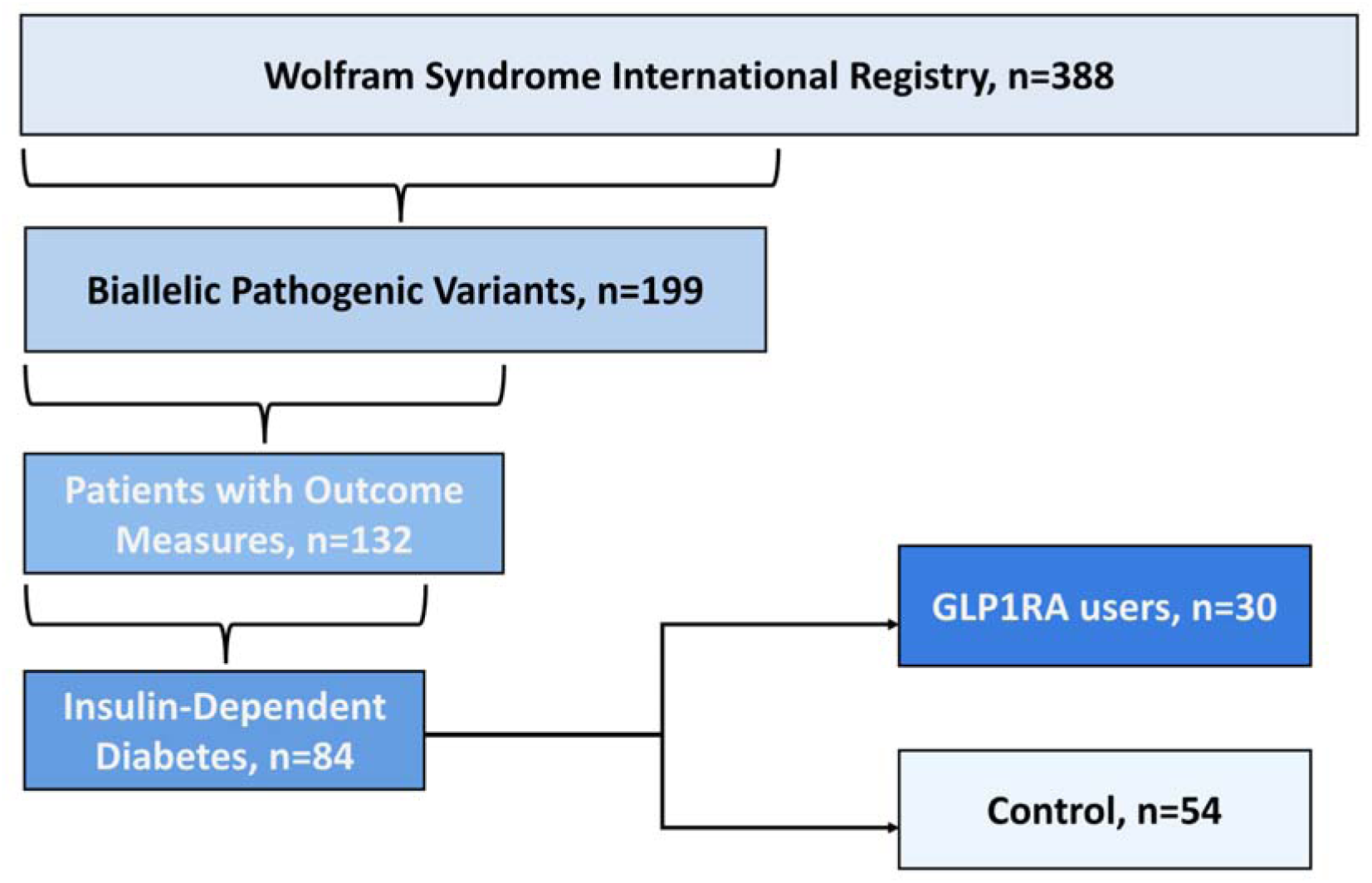
Study population flow diagram. Starting from the Wolfram Syndrome International Registry and Clinical Study (n =388), participants were sequentially filtered to those with confirmed biallelic *WFS1* variants (n = 199), outcome measurements (n = 132), and insulin-dependent diabetes mellitus (n = 84). Of 84 eligible participants, 30 (35.7%) had received GLP-1 RA therapy and 54 (64.3%) remained on insulin alone.

All participants were required to have documented age at Wolfram syndrome diagnosis and age at diabetes mellitus diagnosis. Where applicable, age at diagnosis of optic atrophy, hearing loss, and arginine vasopressin deficiency was also recorded.

Twelve participants had also enrolled in another interventional study in Wolfram syndrome. For these participants, only data collected prior to enrollment in the other interventional study were included.

### Clinical Variables

Data were collected retrospectively through structured chart review and telephone interview. Phone interviews were conducted to verify existing information, capture missing data points, and obtain additional information not available in medical records. Standardized data collection included demographic variables (age, biological sex) and clinical variables.

A genetic severity score was calculated for all participants based on their biallelic *WFS1* variants, ranging from 1 (least severe) to 6 (most severe), based on variant type (in-frame vs. out-of-frame) and location relative to transmembrane domains (https://doi.org/10.64898/2026.03.24.26349216).

Primary endpoints included GLP-1 RA use (current or prior), duration of use, and within-participant changes in BMI, glycemic control, and visual parameters. Secondary endpoints included rationale for initiation, specific agents used, adverse effects, and reasons for discontinuation.

For participants who had taken a GLP-1 RA, outcome measures were analyzed using a pre–post design within two time windows: (1) one year before to one year after GLP-1 RA initiation (Table 2), and (2) one year before to two years after initiation (Table 3). The pre-GLP-1 RA value was the measurement closest to the start date; the post-GLP-1 RA value was the last measurement within the defined window while the participant was actively on the medication. Values obtained after discontinuation were excluded unless the participant had restarted a GLP-1 RA. The following measures were obtained: Body mass index (BMI) (kg/m²); HbA1c (%), C-peptide, fasting glucose, and C-peptide index; best-corrected visual acuity converted from Snellen to LogMAR; and retinal nerve fiber layer (RNFL) thickness (μm) by optical coherence tomography (OCT).

**Table 1.**
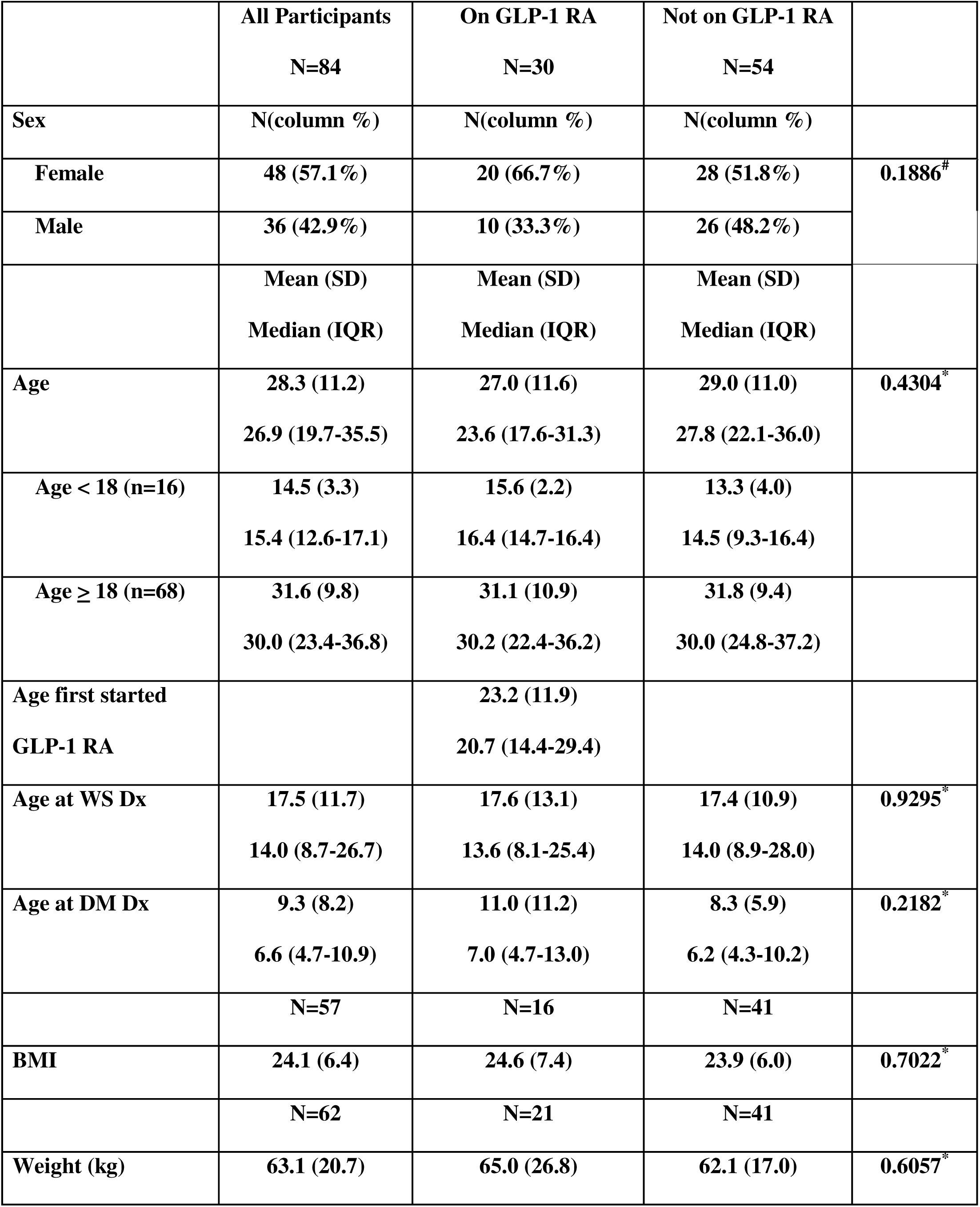
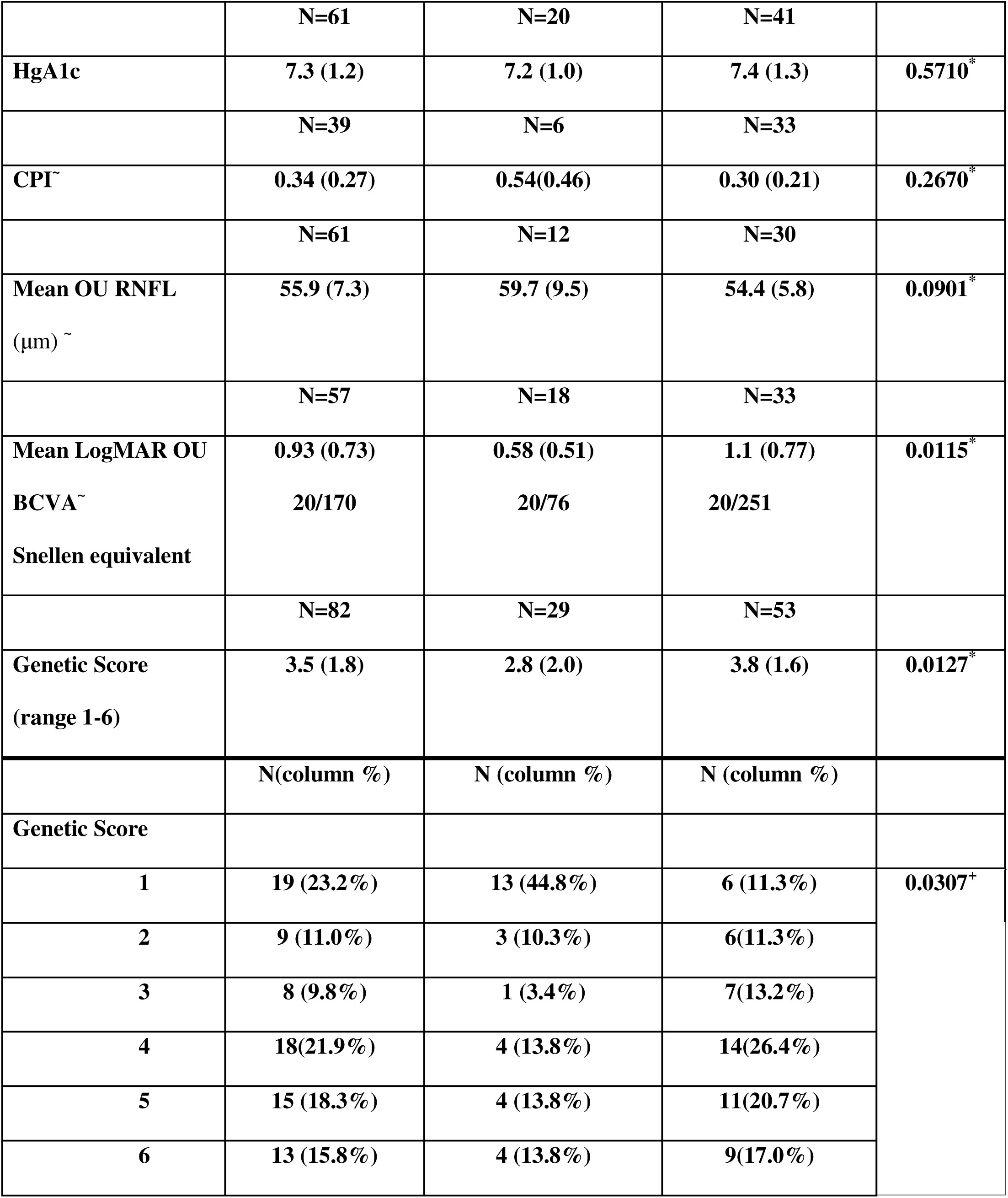
Demographics and clinical characteristics. BCVA = best-corrected visual acuity; BMI = body mass index; CPI = C-peptide index; DM = diabetes mellitus; Dx = diagnosis; GLP-1 RA = GLP-1 receptor agonists; HgA1c = hemoglobin A1c; IQR = interquartile range (1^st^ quartile to the 3^rd^ quartile); OU = both eyes; RNFL = retinal nerve fiber layer; SD = standard deviation; WS = Wolfram syndrome. *p-value from t-test; ^#^p-value from Chi-square test; ^+^p-value from Fisher’s exact test.

**Table 2.**
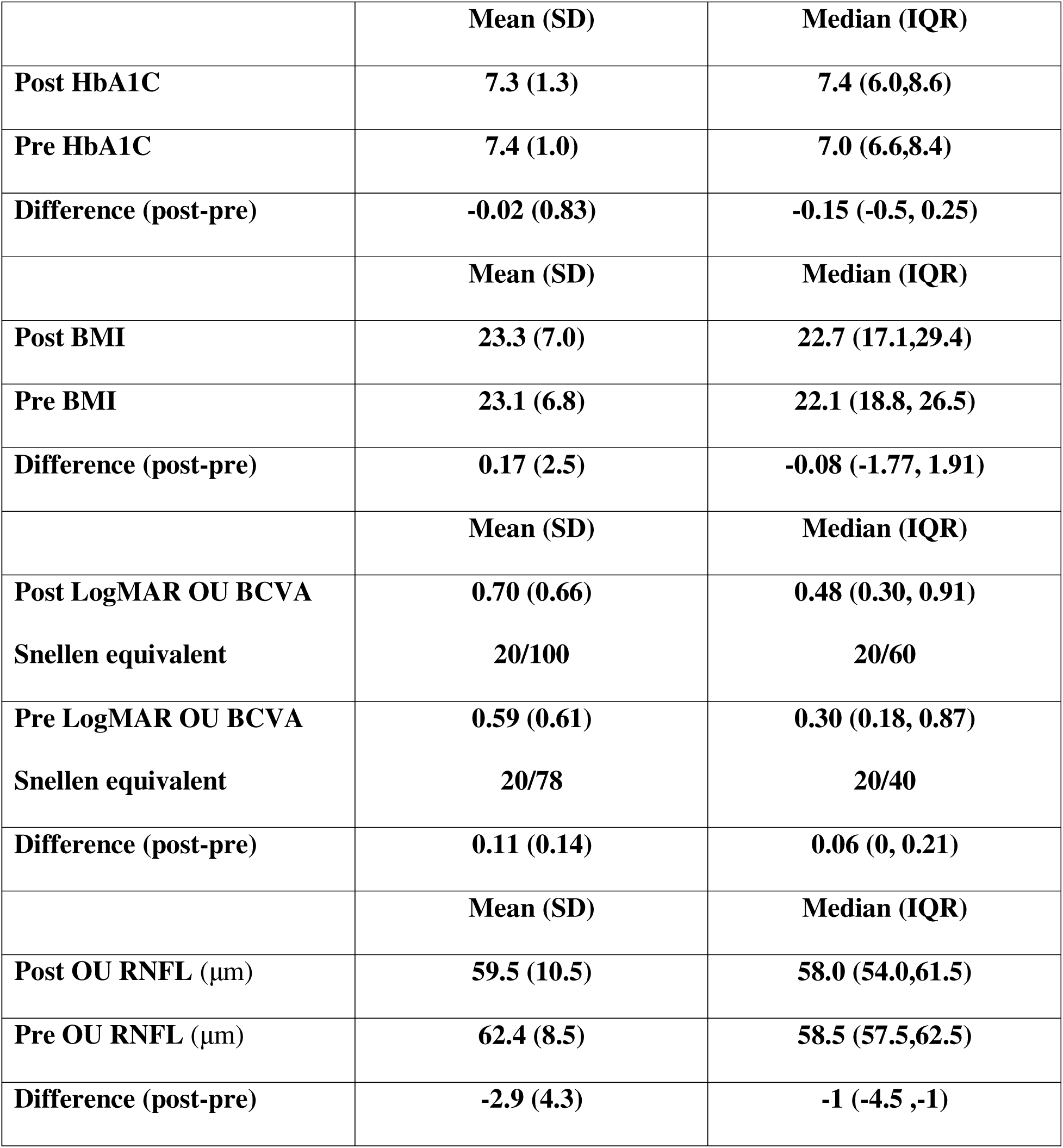
Statistical Analysis Done One Year Before and One Year After Starting GLP-1 RA Medication. BCVA = best-corrected visual acuity; BMI = body mass index; HgA1c = hemoglobin A1c; IQR = interquartile range of the 25^th^ percentile to the 75^th^ percentile; OU = both eyes; RNFL = retinal nerve fiber layer; SD = standard deviation

**Table 3.** Statistical Analysis Done One Year Before and Two Years After Starting GLP-1 RA Medication. BCVA = best-corrected visual acuity; BMI = body mass index; HgA1c = hemoglobin A1c; IQR = interquartile range of the 25th percentile to the 75th percentile; OU = both eyes; RNFL = retinal nerve fiber layer; SD = standard deviation

|  | Mean (SD) | Median (IQR) |
| --- | --- | --- |
| **Post HbA1C** | **7.3 (1.2)** | **7.6 (6.2,8.5)** |
| **Pre HbA1C** | **7.2 (1.0)** | **6.9 (6.5,8.3)** |
| **Difference (post-pre)** | **0.14 (0.78)** | **-0.10 (-0.3, 0.70)** |
|  | Mean (SD) | Median (IQR) |
| **Post BMI** | **23.97 (8.8)** | **24.1 (17.1,25.7)** |
| **Pre BMI** | **23.1 (6.8)** | **22.1 (18.8, 26.5)** |
| **Difference (post-pre)** | **0.84 (4.4)** | **-1.44 (-0.08, 1.70)** |
|  | Mean (SD) | Median (IQR) |
| **Post LogMAR OU BCVA** | **0.71 (0.63)** | **0.48 (0.30, 0.91)** |
| **Snellen equivalent** | **20/102** | **20/60** |
| **Pre LogMAR OU BCVA** | **0.59 (0.61)** | **0.30 (0.18, 0.87)** |
| **Snellen equivalent** | **20/78** | **20/40** |
| **Difference (post-pre)** | **0.12 (0.15)** | **0.16 (0, 0.21)** |
|  | Mean (SD) | Median (IQR) |
| **Post OU RNFL (μm)** | **61.0 (10.1)** | **61.5 (57.0, 62.5)** |
| **Pre OU RNFL(μm)** | **62.4 (8.5)** | **58.5 (57.5,62.5)** |
| **Difference (post-pre)** | **-1.4 (5.5)** | **-1 (-1.5 ,-1)** |

#### Statistical Analysis

Descriptive statistics are reported as mean (SD) and median (IQR). For descriptive variables, we used two-sample t-tests or Fisher’s exact/chi-square tests to compare across groups. Within-participant pre–post changes associated with GLP-1 RA use were evaluated using paired t-tests. Statistical significance was defined by p < 0.05. All analyses were performed using R (version 4.x) and SAS Software for Windows (v9.4).

## Results

Of the 84 participants with insulin-dependent diabetes mellitus who met inclusion criteria, 30 (35.7%) had previously used or were currently using a GLP-1 receptor agonist in addition to insulin therapy, whereas 54 (64.3%) were treated with insulin alone. The cohort was predominantly female (57.1% overall), with a higher proportion in the GLP-1 RA group compared to the non-GLP-1 RA group (66.7% vs. 51.8%, p = 0.18). Mean age was similar between groups (27.0 vs. 29.0 years, p = 0.43). Participants were diagnosed with diabetes mellitus at a mean age of 9.3 years, substantially earlier than their formal diagnosis of Wolfram syndrome at a mean age of 17.5 years.

Participants who received GLP-1 RAs had lower genetic severity scores (mean 2.8 vs. 3.8; p = 0.01) and better baseline best-corrected visual acuity compared to those not receiving GLP-1 RAs (mean LogMAR 0.58 vs. 1.07; p = 0.01), suggesting that patients with milder disease were more likely to receive this adjunctive therapy (Table 1). BMI, weight, HbA1c, and C-peptide index did not differ between groups at baseline (p>.05).

Eight of the 30 GLP-1 RA-treated participants initiated therapy before age 18 years (mean age = 11.6 years; SD = 1.97; median = 11.4 years, IQR = 10.3–12.5). The remaining 22 initiated GLP-1 RA therapy at age 18 or older (mean 27.5 years; SD 11.1; median 24.6 years, IQR 19.0–33.8). The mean duration of GLP-1 RA use was 729.7 days (SD 883.3; median 274.0 days, IQR 68–1452).

### GLP-1 RA Agents and Rationale for Initiation

Semaglutide (n = 14 total, including 2 as second agent) and liraglutide (n = 13) were the most frequently prescribed agents; dulaglutide and tirzepatide were each used by 6 participants (Table 5). The earliest GLP-1 RA initiation in the dataset was in 2019. The most common rationale for initiation was improvement of glycemic control (n = 18), followed by physician recommendation (n = 15), participant-driven research (n = 7), weight management (n = 6), and interest in potential benefits for vision (n = 6) or neurological function (n = 2). Rationale for initiation also included why participants chose the GLP-1 RA agent that they were on. Reasons include participant belief that tirzepatide has better neurological benefits over other GLP-1 RAs (n = 2) and insurance limitations where semaglutide was the only agent insurance companies would cover (n = 3). There were also a few patients who initially began taking liraglutide to participate in a clinical study on the use of liraglutide in Wolfram syndrome patients (n = 5). This study is unrelated to the aforementioned interventional study and all available data for these participants was used for analysis. Reasons for initiation were not mutually exclusive (Table 4).

**Table 4.** Rationale for GLP-1 RA Initiation.

| Rationale | Number of Participants |
| --- | --- |
| Diabetes/Glucose | 18 |
| Medical Advice | 15 |
| External Research | 7 |
| Weight Management | 6 |
| GLP-1 RA Study Trial | 5 |
| Vision | 6 |
| Managing Wolfram syndrome | 5 |
| Insurance Limitations | 3 |
| Participant Interest | 7 |
| Neurological Benefits | 2 |

**Table 5.** Type of GLP-1 RA (First and Second) Taken.

| Type of GLP-1 | GLP-1 RA #1 | GLP-1 RA #2 | Total Number of<br>Participants |
| --- | --- | --- | --- |
| Liraglutide | 13 | 0 | 13 |
| Semaglutide | 12 | 2 | 14 |
| Dulaglutide | 5 | 1 | 6 |
| Tirzepatide | 0 | 6 | 6 |

### Metabolic and Glycemic Outcomes

Within-participant pre–post analyses were conducted for those with available paired data. Baseline HbA1c was relatively well-controlled (mean ∼7.3–7.4%). At one year (Table 2), 12 participants had both pre- and post-HbA1c values. The mean change was −0.02% (SD 0.83; 95% CI −0.56 to 0.51; p = 0.91). At two years (Table 3), 14 participants had paired HbA1c data; the mean change was +0.14% (95% CI −0.31 to 0.59; p = 0.50).

Baseline BMI was normal across the cohort (mean = 24.1, SD=6.4) and was stable at both time points. At one year (n = 13), the mean change was +0.17 kg/m² (SD 2.5; 95% CI −1.32 to 1.66; p = 0.80). At two years (n = 13), the mean change was +0.84 kg/m² (SD 4.42; 95% CI −1.84 to 3.51; p = 0.50). C-peptide index could not be robustly analyzed due to insufficient paired data (only 2 participants with pre-post values).

### Visual Outcomes

Nine participants had paired LogMAR of best-corrected visual acuity data within one year. The mean change (post − pre) was +0.11 LogMAR (SD 0.14; 95% CI −0.002 to 0.22; p = 0.05). At two years, the mean change was +0.12 (SD 0.15; 95% CI 0.006–0.24; p = 0.04), reaching statistical significance and indicating continued visual decline consistent with underlying disease progression (Table 2, Table 3).

Five participants had paired RNFL data. RNFL thickness showed a modest non-significant decline at one year (mean change −2.9 μm; 95% CI −8.2 to 2.4; p = 0.20) and two years (mean change −1.4 μm; 95% CI −8.2 to 5.0; p = 0.59).

### Adverse Effects and Discontinuation

Seventeen of 30 participants (56.7%) reported adverse effects. Gastrointestinal symptoms were the most prevalent, reported by 15 participants (50%), including nausea (n = 6), abdominal pain (n = 5), reduced appetite (n = 5), diarrhea/other GI symptoms (n = 7), and undesired weight loss (n = 2). One participant reported a growth-related side effect. No participants reported vision-related adverse effects (Table 6).

**Table 6.** Adverse Effects of GLP-1 RA.

| Adverse Effect | Number of Participants |
| --- | --- |
| Gastrointestinal (GI) | 15 |
| Digestive | 4 |
| Nausea | 6 |
| Appetite | 5 |
| Weight Loss | 2 |
| Abdominal Pain | 5 |
| Other GI-related | 7 |
| Vision | 0 |
| Growth- Related | 1 |
| Other | 4 |

Thirteen participants discontinued GLP-1 RA therapy (Table 7). The most common reasons were adverse side effects (n = 13, predominantly gastrointestinal), lack of perceived effect (n = 5), enrollment in the clinical trial for a different drug (n = 3), and insurance/pharmacy barriers (n = 2). Two participants discontinued within 10 days of initiation due to severe gastrointestinal intolerance.

**Table 7.** Reasons for Termination of GLP-1 RA.

| Termination Reason | Number of Participants |
| --- | --- |
| Adverse Side Effects | 13 |
| Not Helping/No Effect | 5 |
| Participation in Another Trial | 3 |
| Insurance/Pharmacy | 2 |
| Concern over Potential Side Effect | 1 |
| Want to try another GLP-1 RA | 1 |

## Discussion

This retrospective study represents, to our knowledge, the largest systematic evaluation of GLP-1 RA use in individuals with Wolfram syndrome to date. Among 84 participants with genetically confirmed Wolfram syndrome and insulin-dependent diabetes mellitus enrolled in an international registry, 35.7% had received a GLP-1 RA in addition to insulin therapy, reflecting meaningful clinical adoption of this approach despite a limited evidence base. Our findings provide important clinical context and several key insights.

First, GLP-1 RAs are being used in a substantial and growing proportion of Wolfram syndrome patients. The earliest initiation in our dataset was in 2019, consistent with increased availability of newer formulations and expanding awareness of their potential benefits beyond type 2 diabetes (Drucker, 2018;Nauck et al., 2021). Semaglutide and liraglutide were most frequently prescribed. The major stated rationale was glycemic improvement, and a sizable proportion of patients or clinicians also cited potential vision or neurological benefits, suggesting that preclinical data and anecdotal reports have begun to shape clinical decision-making in this population.

Second, there were no improvements in HbA1c at one or two years. Importantly, baseline glycemic control was already near-optimal (mean HbA1c ∼7.3%), which likely attenuated the ability to detect improvement. In Wolfram syndrome, insulin is the primary treatment for diabetes (Shang et al., 2014;Urano, 2016;Ray et al., 2022); GLP-1 RAs may function as adjunctive agents that modulate residual β cell function and ER stress (Kondo et al., 2018;Gorgogietas et al., 2023). BMI remained stable across both time windows, contrasting with the substantial weight reduction observed in type 2 diabetes trials, likely because Wolfram syndrome participants had normal baseline BMI (Wilding et al., 2021).

Third, visual outcomes did not improve and continued to worsen over the observation period, consistent with ongoing optic nerve atrophy and previous natural history studies (Hoekel et al., 2014;Hoekel et al., 2018;McNeely et al., 2026). RNFL thickness showed a modest non-significant decline. It has been previously noted that RNFL thickness has a bottom-out effect where after the layer thickness reaches a given point, it does not decrease anymore (Mwanza et al., 2015). Additionally, there might also be device specific differences in measurements that limit the reliability and reproducibility of measured RNFL thickness values (Pierro et al., 2012). This suggests that LogMAR of best correct visual acuity may be a better indication of optic nerve atrophy and visual decline across time, especially since there was a significant increase in LogMAR with a nonsignificant decline in RNFL thickness observed in this patient cohort (Yau et al., 2013). A critical limitation of this study is the absence of an untreated comparator group matched for age, disease duration, and genetic severity, which precluded any determination of whether GLP-1 RA therapy slowed, accelerated, or had no effect on the trajectory of vision loss. Further complicating direct comparisons between groups, participants not receiving GLP-1 RAs had significantly higher genetic severity scores and worse baseline visual acuity, reflecting confounding by indication and underlying disease severity.

Fourth, gastrointestinal adverse effects were common, occurring in 56.7% of participants, and led to discontinuation in the majority of those who stopped therapy. This rate is broadly consistent with discontinuation rates reported for GLP-1 RAs in general diabetes and obesity populations (estimated 40–70% at one year) (Gorgojo-Martinez et al., 2022;Rodriguez et al., 2025;Jalleh et al., 2026). Given that autonomic dysfunction and gastroparesis are features of Wolfram syndrome-related neurodegeneration, gastrointestinal intolerance may be amplified in this population. Careful pre-treatment assessment of gastrointestinal symptoms and gradual dose titration may be particularly important in Wolfram syndrome. No vision-related adverse effects were reported.

When interpreted within the context of monogenic diabetes syndromes, our findings are instructive. Unlike certain MODY subtypes (e.g., MODY3 caused by HNF1A variants), in which sulfonylureas produce dramatic glycemic improvement by targeting a specific molecular defect (Pearson et al., 2003), Wolfram syndrome is a progressive ER stress-mediated β-cell failure syndrome with no molecular basis amenable to sulfonylurea therapy (Fonseca et al., 2005;Fonseca et al., 2010;Urano, 2016). In this context, GLP-1 RAs may function primarily as β-cell-protective and ER stress-modulating agents rather than as drivers of large HbA1c reductions. This study has several important limitations. The sample size was modest overall, and the number of participants with paired pre–post measurements was small (9-14 per endpoint), limiting statistical power. The retrospective design introduced missing data, inconsistent measurement intervals, and potential recall bias. Selection bias is likely, as participants engaged in registry research may differ systematically from individuals not enrolled. We were unable to adjust for confounders including age, sex, diabetes duration, insulin regimen, or concurrent investigational therapies. Finally, the absence of a contemporaneous untreated control group limits causal inference regarding disease trajectory.

This study provides a systematic evaluation of GLP-1 RA use in Wolfram syndrome, applied to one of the largest patient cohorts examined to date. The results demonstrate that GLP-1 RAs are already integrated into the care of individuals with Wolfram syndrome and are generally tolerated, though gastrointestinal side effects are frequent. We did not observe significant improvements in glycemic control or visual acuity over one to two years, highlighting the need for prospective, controlled studies. Future work should incorporate standardized endpoints including continuous glucose monitoring metrics, insulin dose requirements, serial C-peptide dynamics, quantitative OCT-based RNFL analysis, and functional visual assessments. Stratification by genetic severity score may clarify whether certain genotypic subgroups derive greater benefit. As some participants noted selection for one agent over another due to differences in perceived impact, it may also be beneficial to compare between various GLP-1 RAs and explore each agent’s neurological and glycemic benefits. As therapeutic strategies targeting ER stress and *WFS1* function continue to advance (Urano, 2016;Abreu and Urano, 2019), rigorous evaluation of GLP-1-based strategies in appropriately powered prospective studies remains a high clinical and scientific priority.

## Author Contributions

LL, AT, and AA contributed equally as co-first authors: conceptualization, data collection, data analysis, and manuscript writing. SN, HR, and ED contributed to data collection and manuscript review. HL, JH, BM, and TH contributed to study design, clinical data acquisition, and manuscript review. FU supervised the project, provided critical conceptual input, secured funding, and approved the final version. All authors contributed to manuscript preparation and are accountable for the integrity of the work.

## Acknowledgments

We sincerely thank the members of the Washington University Wolfram Syndrome and Related Disorders Clinic and Research Team (https://wolframsyndrome.wustl.edu) for their invaluable support. We are especially grateful to all participants in the Wolfram Syndrome and Related Disorders International Registry, Clinical Studies, Longitudinal Study, and Clinical Trials for their time, dedication, and commitment to advancing research in this field.

We also thank Dr. Sarah Gladstone and Mrs. Stephanie Snow Gebel of the Snow Foundation for their encouragement and support in initiating this project. We gratefully acknowledge the generous philanthropic support of the Auerbach Hyman Fund, the Philipp Fund, the Jerome W. Gratenstein Memorial Foundation, the WAVE Fund, the WAV Fund, the Silberman Fund, the Stowe Fund, the Feiock Fund, the Cachia Fund, the Gildenhorn Fund, the Snow Foundation, the Ellie White Foundation for Rare Genetic Disorders, the Unravel Wolfram Syndrome Fund, the Be A Tiger Foundation, and the Eye Hope Foundation. We further acknowledge the support of our international partner organizations: the Ontario Wolfram League, Associazione Gentian Sindrome di Wolfram Italia, Alianza de Familias Afectadas por el Síndrome de Wolfram Spain, Wolfram Heroes Society, Wolfram Syndrome UK, and Association Syndrome de Wolfram France. Their advocacy and dedication to the Wolfram syndrome community are deeply appreciated.

## Ethics statement

The studies involving humans were approved by the Washington University in St. Louis Human Research Protection Office. The studies were conducted in accordance with the local legislation and institutional requirements. Children under age 18 gave informed assent, and parents/guardians gave informed, written consent. Participants 18 or older gave informed, written consent for participation in this study.

## Data Availability Statement

The deidentified data supporting the findings of this study will be made available from the corresponding author, Fumihiko Urano, MD, PhD, upon reasonable request and subject to institutional review and execution of an appropriate data use agreement.

